# Age-Associated Expression of DIMT1 (Dimethyladenosine Transferase) in Human Mesenchymal Stromal Cells: A Candidate for Multi-Gene Aging Signatures

**DOI:** 10.1101/2025.11.16.25340333

**Authors:** James Utley

**Affiliations:** Syndicate Laboratories

## Abstract

**Background:** DIMT1 is a highly conserved methyltransferase that functions as an N_6_,N_6_-dimethyladenosine transferase responsible for modifying two adjacent adenosines in the 18S rRNA. This protein is also a crucial, yet catalytically independent, ribosome maturation factor essential for the biogenesis of the 40S small ribosomal subunit. Given the established roles of ribosome biogenesis and methylation dynamics in biological aging, we investigated the relationship between DIMT1 expression and chronological donor age in Mesenchymal Stromal Cells (MSCs).

**Methods:** Transcriptomic data for DIMT1 from a publicly available human MSC dataset (*n* = 61 donors, age range: 17–84 years) were analyzed. Pearson correlation and simple linear regression were performed to assess the association between normalized gene expression and donor age.

**Results:** A statistically significant positive correlation was identified between DIMT1 expression and donor age (Pearson *r* = 0.4282, *R*^2^ = 0.183, *p* = 0.0006). Linear regression modeling demonstrated that expression increases with advancing age, exhibiting a slope of 0.0045.

**Conclusion:** DIMT1 expression demonstrates a statistically significant positive correlation with donor age in MSCs, though age explains only 18.3% of expression variance. This relationship warrants further investigation of DIMT1 as a component of multi-gene aging signatures. The observed pattern may reflect compensatory responses in ribosome biogenesis or altered methylation dynamics associated with aging, though mechanistic validation is required.

## 1 Introduction

Biological aging is a complex, multifactorial process marked by a progressive decline in cellular and systemic function. Key hallmarks of aging include genomic instability, telomere attrition, altered intercellular communication, and dysregulation of cellular metabolic and biosynthetic pathways (Alves et al., 2012). Identifying molecular markers that accurately reflect biological age and mechanistic shifts remains a primary goal in geriatric and regenerative medicine.

*DIMT1* (DIM1 rRNA Methyltransferase and Ribosome Maturation Factor) is critical to ribosomal integrity. It encodes a methyltransferase that specifically dimethylates two adjacent adenosines (A1818 and A1819 in human) in a conserved hairpin loop near the 3*′*-end of the 18*S* rRNA within the 40*S* small ribosomal subunit (on UniProtKB/Swiss-Prot Summary, 2024; Pulicherla et al., 2009). Beyond its catalytic activity, the *DIMT1* protein is an integral component of the small subunit (SSU) processome (Liberman & et al., 2021), where it participates in pre-rRNA processing steps independently of its methyltransferase function (Liu et al., 2020). The methyl donor for this reaction is S-adenosylmethionine (SAM), placing *DIMT1* directly within the methionine-methylation cycle, a metabolic pathway tightly linked to age-related epigenetic drift (Issitt et al., 2023). Given the essential role of MSCs as a model for cellular senescence and the pivotal function of *DIMT1* in cellular biosynthesis, we hypothesized that its expression level would correlate significantly with chronological donor age.

## 2 Methods

### 2.1 Data Acquisition and Preparation

Transcriptomic data from human Mesenchymal Stromal Cells (MSCs) were utilized for this analysis. The dataset was obtained from a previously published study by Alves et al. (2012) (Alves et al., 2012), which provides normalized gene expression profiles and corresponding chronological donor age metadata. The specific gene of interest was identified by its common name, *DIMT1*, and its unique Ensembl ID, ENSG00000086189. All expression values were used as processed and normalized by the original authors.

### 2.2 Sample Characteristics

The dataset comprised n = 61 human MSC samples from donors ranging in age from 17 to 84 years (mean = 55.23 years, SD = 17.26 years).

### 2.3 Statistical Analysis

The association between the normalized expression level of *DIMT1* and donor chronological age was quantified using the Pearson product-moment correlation coefficient (*r*). Statistical significance was determined using a two-tailed *p*-value. Subsequently, a simple linear regression model was fitted to the data to model the dependency of gene expression (dependent variable) on donor age (independent variable). A significance threshold of *p <* 0.05 was applied for all inferential tests.

**Figure 1.**
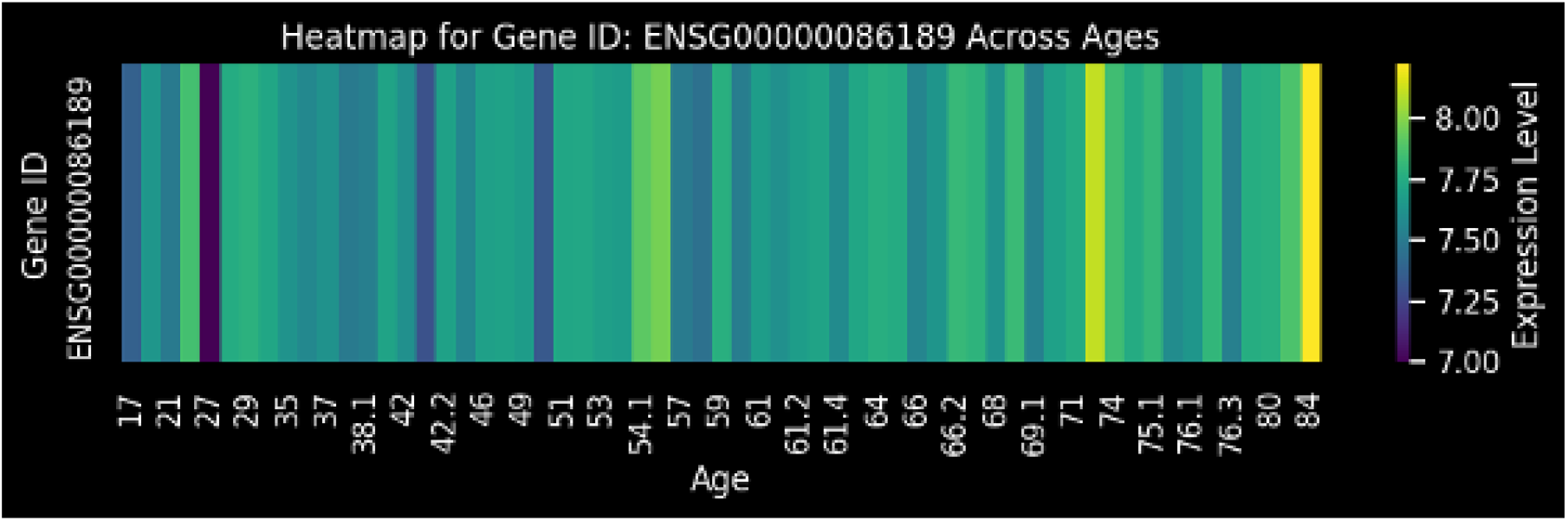
Heatmap Visualizing *DIMT* 1 Gene Expression Level Across Different Ages. The heatmap displays the logscale expression level of the *DIMT* 1 gene (ENSG00000086189) for each individual, ordered by age (x-axis). The color intensity corresponds to the expression level, as indicated by the scale bar on the right: darker purple/blue represents lower expression (around 7.00), and brighter yellow/green represents higher expression (up to 8.00). The pattern suggests a general trend of increasing expression in older individuals (right side of the plot), consistent with the positive correlation observed in the linear regression analysis (Figure 1).

## 3 Results

### 3.1 Descriptive Expression Profile

Analysis of *DIMT1* expression across all donor samples yielded a mean expression level of 7.6742 (standard deviation, *SD* = 0.1820). The observed expression range spanned from a minimum of 6.9992 to a maximum of 8.2232 (Table 1).

**Table 1:** Statistical Summary of *DIMT1* Expression and Age Correlation in MSCs.

| Metric Type | Statistic | Value | Interpretation |
| --- | --- | --- | --- |
| Descriptive | Mean Expression | 7.6742 | Average log-scale expression |
|  | Standard Deviation | 0.1820 |  |
|  | Minimum Expression | 6.9992 |  |
|  | Maximum Expression | 8.2232 |  |
| Inferential | Pearson Correlation ($r$) | 0.4282 | Significant positive linear relationship |
| | **$p$-value** | **0.0006** | Highly statistically significant ($p < 0.05$) |
|  | Linear Regression Slope | 0.0045 | Expression increase per year of age |
|  | Linear Regression Intercept | 7.4249 | Predicted expression at age zero |
|  | Direction | Positive | Upregulation with age |
| | $R^2$ (Coefficient of Determination) | 0.183 | Age explains 18.3% of expression variance |

### 3.2 Age-Dependent Correlation

A statistically significant positive correlation was identified between the expression level of *DIMT1* and chronological donor age (Pearson *r* = 0.4282, *p* = 0.0006) The coefficient of determination (R^2^ = 0.183) indicates that chronological age explains approximately 18.3% of the variance in DIMT1 expression levels. This result confirms the initial hypothesis, indicating a clear upregulation of *DIMT1* transcription with increasing donor age. The analysis included 61 MSC samples from donors with ages ranging from 17 to 84 years (mean age: 55.23 *±* 17.26 years).

**Figure 2.**
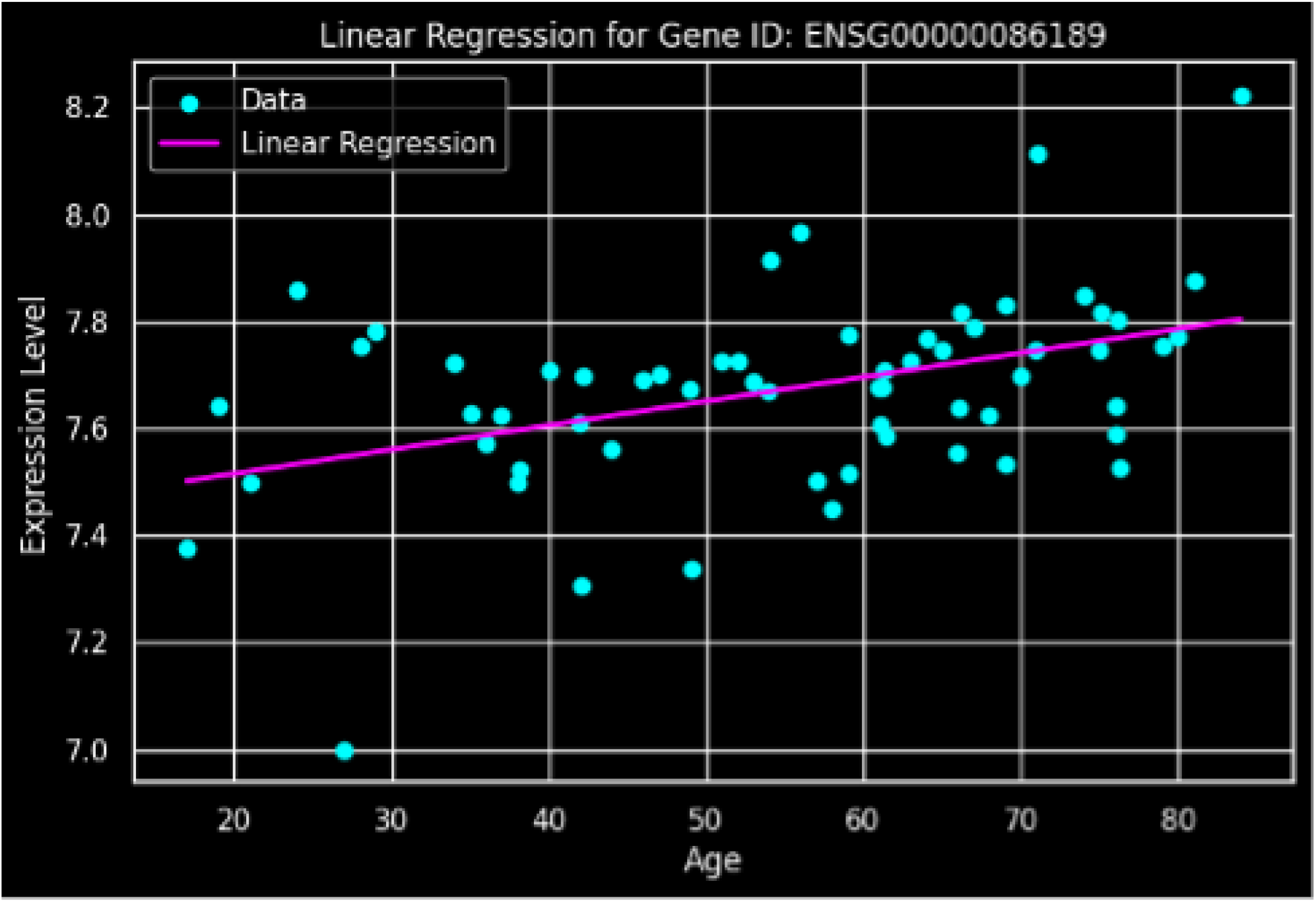
Linear Regression Analysis of *DIMT* 1 Gene Expression Level vs. Age in Mesenchymal Stem Cells (MSCs). The plot displays the relationship between the log-scale expression level of *DIMT* 1 (y-axis) and the age of the individual (x-axis). Each cyan point represents the data for a single individual. The magenta line shows the fitted linear regression model: Expression Level = 7.4249 + 0.0045 × *Age* (as determined by the linear regression intercept and slope in Table 1). The positive slope and the statistically significant positive Pearson correlation (*r* = 0.4282, *p <* 0.05) indicate that *DIMT* 1 expression significantly increases (is upregulated) with age in MSCs.

### 3.3 Linear Regression Modeling

The simple linear regression model provided a quantitative measure of this age-dependent increase. The fitted regression equation is:

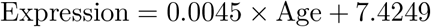

The positive slope coefficient (*β* = 0.0045) signifies an average increase of 0.0045 arbitrary units in *DIMT1* expression for every additional year of life of the MSC donor. The comprehensive statistical summary is provided in Table 1.

## 4 Discussion

The identification of a statistically significant, moderate positive correlation between the expression of DIMT1 and chronological age in human MSCs (r = 0.4282, R2 = 0.183) suggests this gene warrants further investigation as a potential biomarker of biological aging, though age explains only 18.3% of the observed variance in expression.The functional annotation of *DIMT1* as a methyltransferase and ribosome maturation factor suggests that the observed transcriptional upregulation may be mechanistically linked to fundamental age-related processes.

The dual role of *DIMT1* is highly relevant to aging theory. Firstly, its involvement in *N* ^6^, *N* ^6^-dimethyladenosine modification of 18*S* rRNA connects it to the fidelity and efficiency of translation, which is often compromised in senescence. Secondly, its placement in the SAM-dependent methylation cycle links it to global epigenetic stability. An increase in *DIMT1* expression could represent:

1. A Compensatory Mechanism: An attempt by aging cells to maintain ribosomal biogenesis efficiency despite accumulating cellular damage, or to bolster methylation capacity in the face of declining methionine cycle flux.
2. A Maladaptive Response: A shift in methylation stoichiometry leading to aberrant gene regulation or an imbalance in the SSU processome that contributes to age-related cellular dysfunction.

The fact that *DIMT1* is essential for ribosome biogenesis even without its catalytic activity underscores its role as a scaffold or structural component of the SSU processome (Liu et al., 2020). Supporting this hypothesis, studies in C. elegans demonstrated that modulation of the *dimt-1* ortholog significantly regulates organismal lifespan and stress resistance via selective translation of mRNA transcripts, linking this enzyme to major longevity pathways such as DAF-9 signaling (Rothi et al., 2025). The age-related increase in its transcript may be necessary to support the complex, nuclearly-localized assembly of the 40*S* subunit under conditions of age-related stress.(Rothi et al., 2025)

### 4.1 Limitations and Future Directions

The reliance on a single, albeit robust, public transcriptomic dataset limits the generalizability of these findings. Future studies should focus on external validation in diverse human tissues and cell types. Crucially, mechanistic studies employing CRISPR/Cas9 knockdown or overexpression of *DIMT1* are required to determine if its dysregulation is a cause or merely a consequence of the aging phenotype. Furthermore, assessing the activity of the DIMT1 protein not just its transcript level will be necessary to fully elucidate its role in age-related ribosomal and epigenetic changes. It is important to note that the mechanistic pathway illustrated in Figure 3, linking oxidative stress to DIMT1 upregulation, represents a hypothetical model inferred from the literature and the observed correlation. The present study does not include direct measurements of oxidative stress markers, DIMT1 protein levels, methylation activity, or ribosome biogenesis efficiency. The proposed mechanism requires experimental validation through:

**Figure 3.**
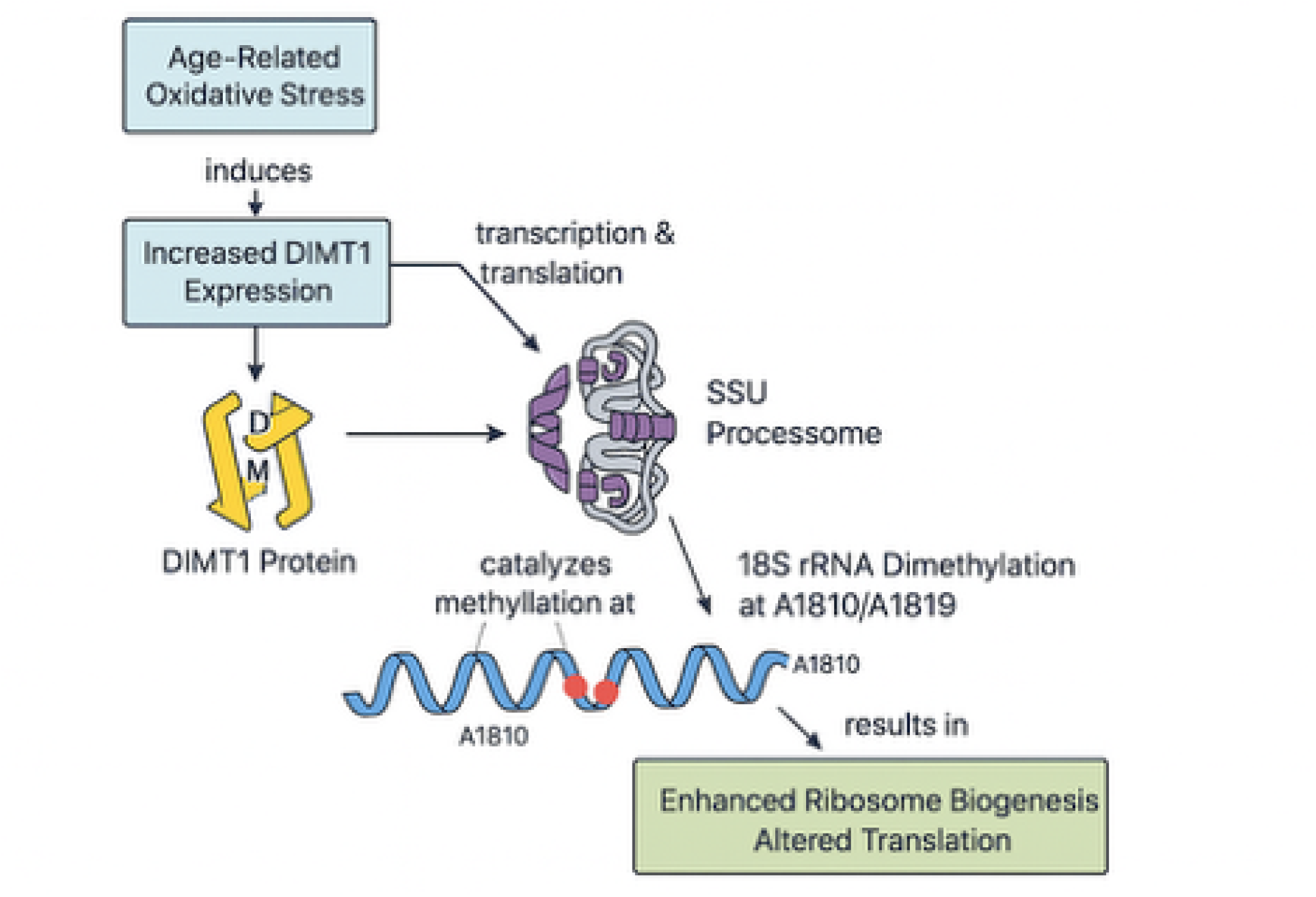
Hypothetical Model: Proposed Mechanism of *DIMT* 1-mediated Ribosome Biogenesis Regulation in Response to Age-Related Oxidative Stress. Age-related oxidative stress induces the expression of the *DIMT* 1 gene. The resulting DIMT1 protein (Dimethyladenosine Transferase 1) is recruited to the SSU (Small Subunit) Processome. Here, DIMT1 catalyzes the methylation of the 18S ribosomal RNA at the A1818/A1819 positions. This methylation step is critical for 18*S* rRNA maturation and ultimately results in enhanced ribosome biogenesis and altered translation, linking cellular aging signals to protein synthesis capacity.

- Measurement of oxidative stress markers in age-stratified MSC samples
- Correlation of DIMT1 transcript with protein abundance and enzymatic activity
- Direct quantification of 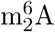 modifications at positions 1818/1819 in 18S rRNA
- Functional assessment of ribosome biogenesis rates in high vs. low DIMT1 expressors

No adjustments were made for potential confounding variables such as donor sex, variation in quality of tissue source, consistency in culture passage number and cell quality, as the primary objective was to assess the bivariate relationship between age and DIMT1 expression. The limitations of this approach will need to be investigated in future research.

### 4.2 Interpretation of Effect Size

While the correlation between DIMT1 expression and age is statistically robust, the modest coefficient of determination (*R*^2^ = 0.183) indicates that chronological age alone explains less than one-fifth of DIMT1 expression variability. The remaining 81.7% of variance likely reflects biological factors including individual genetic variation, environmental exposures, replicative history of the MSC cultures, and technical variation. This effect size is consistent with single-gene biomarkers in complex biological processes but suggests DIMT1 would be most informative when integrated into multi-gene predictive models of biological age.

## 5 Conclusion

DIMT1 expression demonstrates a statistically significant, moderate positive correlation with chronological donor age in Mesenchymal Stromal Cells (*r* = 0.4282, *R*^2^ = 0.183). While this relationship is reproducible and mechanistically plausible given DIMT1’s role in ribosome biogenesis and methylation, age explains less than one-fifth of the observed expression variance. These results support continued investigation of DIMT1 in the context of multi-gene aging signatures and warrant validation in independent cohorts before clinical biomarker utility can be established. The modest effect size suggests DIMT1 may be more valuable as part of a composite biomarker panel rather than as a standalone aging indicator.

## Data Availability

All data produced are available online at https://www.stemformatics.org/datasets/view?id=6183

## Notes

### Competing Interest Statement

The authors have declared no competing interest.

### Funding Statement

No funding was provided for this study

### Author Declarations

https://www.stemformatics.org/datasets/view?id=6183

## References

Alves, H., van Ginkel, J., Groen, N., Hulsman, M., Mentink, A., Reinders, M., van Blitterswijk, C., & de Boer, J. (2012). A mesenchymal stromal cell gene signature for donor age. PLoS One, 7 (8), e42908. 10.1371/journal.pone.0042908

Issitt, T., Mason, A. S., & Sweeney, S. T. (2023). Cellular response to starvation provides biomarkers for breast cancer through volatile metabolites linked to methylation and methionine metabolism [Doctoral dissertation, White Rose eTheses Online]. https://etheses.whiterose.ac.uk/id/eprint/34381/1/Issitt_204032380_CorrectedThesis_Clean.pdf

Liberman, N., & et al. (2021). Intergenerational hormesis is regulated by heritable 18s rrna methylation. bioRxiv. 10.1101/2021.09.27.461965

Liu, G., Peng, X., Cai, Y., & et al. (2020). Structural and catalytic roles of the human 18 s rrna methyltransferases dimt1 in ribosome assembly and translation. Nucleic Acids Res, 48 (15), 8617–8630. 10.1093/nar/gkaa641

on UniProtKB/Swiss-Prot Summary, B. (2024). N 6, N 6-dimethyladenosine transferase activity of dimt1 in 18s rrna. Unpublished data or Database Annotation.

Pulicherla, N., Abba, T. P., Bhaumik, A., & Sanyal, S. (2009). Structural and functional divergence within the dim1/ksga family of rrna methyltransferases. Journal of Molecular Biology, 391 (5), 884–893. 10.1016/J.JMB.2009.06.015

Rothi, M. H., Sarkar, G. C., Al Haddad, J., & et al. (2025). The 18s rrna methyltransferase dimt-1 regulates lifespan in the germline later in life. Nature Communications, 16 (1). 10.1038/s41467-025-62323-7

